# Self-testing knowledge and beliefs on HIV self-testing use in central Uganda

**DOI:** 10.1101/2024.01.12.24301209

**Authors:** Gerald Mukisa Nsereko, Loyce Kiiza Kobusingye, Khamisi Musanje, Joanita Nangendo, Simon Nantamu, Martin Mabunda Baluku

**Affiliations:** Department of Educational, Social and Organizational Psychology, Makerere University, Kampala, Uganda; Clinical Epidemiology Unit, College of Health Sciences, Makerere University, Kampala, Uganda

## Abstract

The government of Uganda and its implementing partners have made significant investments in HIV self-testing (HST) services to reduce clinic-related barriers to HIV testing. However, although HSTs have been around for a reasonable time, their uptake, especially among unskilled working individuals, has remained suboptimal, which threatens the efforts to achieve the 95-95-95 UNAIDS goals to end HIV by 2030.

This study set out to examine whether knowledge and individual beliefs about self-testing influence the use of HIV self-tests among causal workers in Kampala. This cross-sectional study was conducted among 453 participants selected via systematic random sampling from Darling Uganda Ltd. in Wakiso district between July and August 2023. Standardized questionnaires were used, and the analysis was performed using Pearson Correlations and Linear Regression in SPSS.

The findings indicate that HIV self-testing (HST) knowledge was positively related to HST use (r=.387, p≤.01), perceived susceptibility (r=.212, p≤.01), perceived benefits (r=.152, p≤.01), and perceived barriers (r=.101, p≤.05). Individual beliefs, that is, perceived susceptibility (r=.355, p≤.01), perceived benefits (r=.487, p≤.01), and perceived barriers (r=.148, p≤.01), were significantly related to HST use. Perceived benefits were the best predictor of HST use among this population (*β*=.442, *p*=.000). Therefore, (44.2%) of HST use was attributed to the benefits attached to the service.

Therefore, increasing awareness regarding the benefits of HST among such populations increases the chances of HIV testing, particularly among men. This can pave the way for the 95-95-95 goal by 2030 of UNAIDS.

## Introduction

HIV remains a major global public health challenge. By 2022 [1], approximately 40 million people were estimated to be living with HIV, of which 25.7 million people live in sub-Saharan Africa [1]. In Uganda alone, about 1.5 million people are estimated to live with HIV [1]. In 2022, of the 1.5 million HIV infections in Uganda, 5.8% occurred among people aged 15-49 years and above who form the largest percentage of the national workforce [2]. This creates a need to intensify interventions to reduce HIV infection in this sub-population. To reduce the burden of HIV and possibly end it by 2030, the UNAIDS proposed ambitious 95-95-95 targets in 2020[1]. Uganda has made strides toward the 95-95-95 HIV UNAIDS target [2, 3].

Provider stigma, that is, fear of being seen testing at the clinic, distance to health centres (in some rural areas), financial constraints, and access to testing sites, have been identified as the main barriers to HIV testing [5]. Incidentally, most of these are common to clinic-based testing, hence a need to promote other HIV testing methods [6]. HST services are increasingly gaining traction as important alternatives to counteract barriers surrounding clinic-based testing [7]. They are considered private and convenient ways to help people who do not know their status [7]. By employing either oral swab or finger-prick self-test kits, the person can carry out this action wherever it’s most convenient for them; at home, at work, or elsewhere [8, 9, 10].

The increase in infections among adults has been attributed to limited testing, for example, in 2018, approximately 60% of adults in Uganda were reluctant to test for HIV given the stigma surrounding a positive HIV test result [2, 11]. Knowledge of one’s HIV status is considered the first step in the HIV care cascade and an entry point for reducing HIV infections [12, 13]. In this regard, the Uganda Government has launched several community-based campaigns to create awareness and encourage voluntary testing among the population [2]. While community-targeted strategies are important in promoting public health initiatives, certain contexts, like workplaces, seem to miss out on such initiatives, yet adults spend more time at the workplace than in communities [14]. Due to the hectic schedules, tight deadlines, and occasionally inconvenient and time-consuming commutes to pharmacies or medical facilities from work, this population finds it hard to do testing for HIV [4].

HIV self-testing Kits are available for free in public health centers and at low cost in pharmacies to increase accessibility. However, test accessibility does not guarantee successful coverage and use [15]. Perceived susceptibility, (individual knowledge about the tests and attitudes/beliefs), perceived benefits and perceived barriers towards actual usage, maybe be better understood through obtaining insights into the targeted potential users [16]. The Health Belief Model (HBM) proposes that for individuals to change their behavior towards a health intervention, they are influenced by both modifying factors and individual beliefs [17]. This study aimed to test whether HST knowledge and individual beliefs predict use of HST services among workers through the health beliefs model lens.

## Methods

### Study design and setting

We conducted a cross-sectional survey between June and September 2023 to gather information from many respondents in a short period of time and in the most effective way [18]. The study is part of a bigger study titled “Effect of a workplace based HIVST intervention on HIVST use among unskilled workers in Wakiso”. It was undertaken in Darling Uganda Ltd, a manufacturing company located on Jinja Road, Namanve Industrial area. The organization is a component of the Darling Group, which conducts business in 18 nations in Eastern, Western, Southern, and Central regions of Africa, producing premium hair extensions produced from premium Japanese fiber for both the domestic and foreign markets. The company has a little over 3,000 unskilled workers who support the entire production process coming from several units like packing, sorting, lifting, sealing among others. These require a sizable number of human resource due to the high demand of hair extensions in the Ugandan market and beyond. Majority of the employees have attained lower high school education with basic life skills that help them perform tasks given to them in Darling Uganda.

### Study population

Participants were enrolled into the study during the month of June 2023 if they met the following eligibility criteria; aged 18 years and above and willing to offer a written informed consent to join the study. Participants were excluded if they showed disinterest in participating.

### Ethics

The study was approved by the Makerere University School of Medicine Research Ethics Committee; Mak-SOMREC-2022-515 and received administrative clearance from the participating study site (Darling Uganda Limited). All prospective participants gave written consent prior to joining the study.

### Sampling and sample size

The modified Kish Leslie formula (N = (Z2α/2P(1-P))/ d2/*DE) (21) was used to determine the required sample size. A 95% confidence interval equivalent to 1.96 (Z2α), assuming a design effect (DE) = 2, 5% tolerable random error (d), 15% probability of not using the HST and (P) 85% sample proportion that uses HST [19] informed the study. An estimated minimum sample size of 425 unskilled workers both male and female was arrived at. However, 453 was the total of participants who agreed to engage in the study.

Systematic random sampling was used. Here the participants were listed and numbered according to the sampling frame, the population (3000) was divided by the required sample size of 453. An interval fraction of (7) was used to select elements for inclusion into the study depending on the inclusion and exclusion criteria.

### Data collection

The study adapted self-administered closed-ended questionnaires; data was gathered on a 5-point Likert scale ranging from 1 (strongly disagree) to 5 (strongly agree). HST knowledge was measured using HIV self-testing knowledge questionnaire HSTKQ (Gumede & Sibiya, 2018). The HSTKQ is a 15-item tool with a reliability of (α = .811). Individual beliefs were measured with the HIV Testing Belief Scale (HTBS) developed by Alemayehu (2015). The HTBS is a 39-item tool designed to measure the HBM constructs. The current study used 3 items for perceived susceptibility with a reliability of (α = .485), 6 items for barriers (α = .718), and 6 items for benefits (α = .631) as they relate to workers’ use of HST. Self-testing use was measured using a nine-item tool for HIV self-testing (used in Senegal) to measure use and acceptability (Lyons et al., 2019) with a reliability index of (α = .861). The tools were translated into Luganda (a local dialect) to help in easy comprehension.

### Data analysis

Data was analysed using descriptives, Pearson Product Moment Correlation Coefficient (PPMCC), and regression. Descriptives of the different categories of background characteristics were presented to show the percent distribution of respondents. Correlational and Linear regression was used to examine HST knowledge, individual beliefs and HST use among unskilled workers in Wakiso.

## Results

Cisgender female respondents were 53% more than the cisgender males at 47%. About 29.2% of the respondents were aged 22-25. Majority of the participants were married 33.8%. Concerning religion, majority were born again Christians at 32.9%. From the study results, 40.6% of the respondents had a high school level of education. Regarding sexual activity, 64.9% reported to having been sexually active in the past 6 months. Table 1 summarizes the demographic information of the study participants.

**Table 1:** Descriptives.

| Variable | Levels | Frequency | Percentage |
| --- | --- | --- | --- |
| Sex | Male | 213 | 47.0 |
|  | Female | 240 | 53.0 |
|  | Total | 453 | 100.0 |
| Age | 18-21 | 105 | 23.4 |
|  | 22-25 | 131 | 29.2 |
|  | 26-29 | 76 | 17.0 |
|  | 30-33 | 58 | 12.9 |
|  | 34-37 | 26 | 5.8 |
|  | 38-41 | 42 | 9.4 |
|  | Above 41 | 10 | 2.2 |
|  | Total | 453 | 100.0 |
| Level of education | Primary | 80 | 18.1 |
|  | Highschool | 180 | 40.6 |
|  | Certificate | 99 | 22.3 |
|  | Diploma | 40 | 9.0 |
|  | Other | 44 | 9.9 |
|  | Total | 453 | 100.0 |

HST knowledge is positively related to perceived susceptibility (*r*=.212, *p*≤.01), perceived benefits (r=.152, p≤.01), perceived barriers (r=.101, p≤.05), and HST use (r=.387, p≤.01). Individual beliefs of perceived susceptibility (r=.355, p≤.01); perceived benefits (r=.487, p≤.01); and perceived barriers (r=.148, p≤.01) are significantly related to HST use.

The regression results in Table 3 indicate that perceived benefits are the biggest predictor of HST use among unskilled workers (*β*=.442, *p*=.000) that is, 44.2% of HST use is attributed to perceived benefits. This is followed by HST knowledge (*β*=.285, *p*=.000), susceptibility (*β*=.140, *p*=.001), and finally barriers (*β*=.048, *p*=.097). HST knowledge, susceptibility, benefits, and barriers predict HST use by 35.4% [Ad *R*^2^=.354, *F* (62.9), *p*=.<001].

**Table 2:** correlation results.

| P | 1 | 2 | 3 | 4 | 5 |
| --- | --- | --- | --- | --- | --- |
| 1. HST Knowledge | 1 |  |  |  |  |
| 2. Perceived Susceptibility | .212** | 1 |  |  |  |
| 3. Perceived Benefits | .152** | .374** | 1 |  |  |
| 4. Perceived Barriers | .101* | .111* | .101* | 1 |  |
| 5. HST Use | .387** | .355** | .487** | .148** | 1 |

\* $p < .05$ , \*\* $p < .01$ , N = 453

**Table 3:**
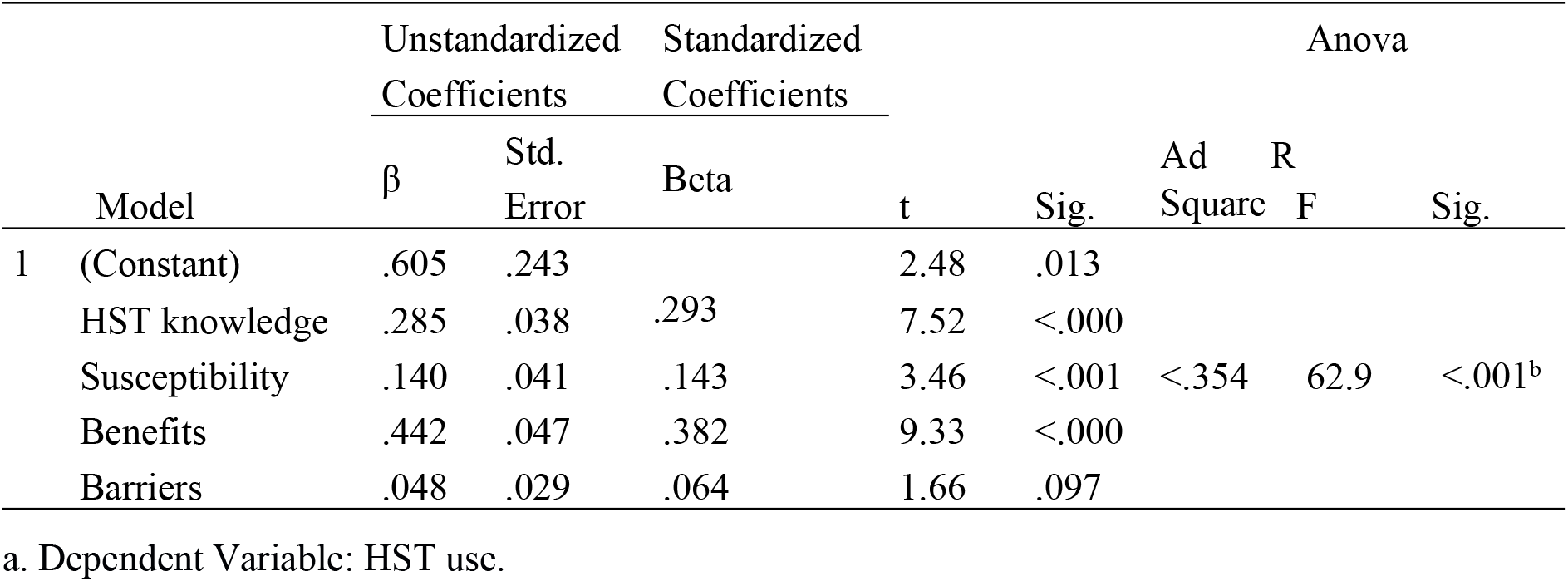
Regression.

## Discussion

The study intended to examine the influence of HST knowledge and individual beliefs on HST use in central Uganda. The findings indicate that HIV self-testing (HST) knowledge was positively related to HST use, perceived susceptibility, perceived benefits, and perceived barriers. Individual beliefs, that is, perceived susceptibility, perceived benefits, and perceived barriers, were significantly related to HST use. Perceived benefits were the best predictor of HST use among this population.

It was hypothesized that HST knowledge significantly relates and predicts HST use. The findings indicate that there exists a positive significant relationship between HST knowledge and HST use nevertheless, it predicts up to 28.5% of HST use among unskilled workers. Employees who possess adequate or basic knowledge about HST such as, the time it takes to get back the results, the accuracy of the results, the interpretation of results, and how HST is administered among others take an extra step and use the HST kit. Additionally, such an employee is likely to be more careful with his/her life/health by ensuring HIV testing comes first before engaging in any transactional or non-transactional sexual behavior which might be risky.

These findings corroborate those of Vara and colleagues who found out that possession of knowledge about HIV richly aids in overcoming fear and denial, that is, people that were initially afraid of HIV and in denial of its existence around them is overcome once they get to learn more about it which then influences their perceptions towards it and contributes to behavior modification [20]. Similarly, being knowledgeable about prevention, treatment, transmission and other facts about HIV, encourages logical safe sex behavior [21]. Therefore, the possibility that the possession of adequate and accurate knowledge is highly correlated to preventive efforts is a powerful motivating factor in most educational projects related to HIV/AIDS. The intention to utilize HST has been proven to be influenced by knowledge [20].

It was hypothesized that HST knowledge is significantly related to individual perceived susceptibility. The findings indicate that there is a significant relationship between HST knowledge and individual perceived susceptibility. An employee who has full information regarding HST is most likely to have high perceived susceptibility simply because being aware of what the test can do and how to use it effectively makes an individual perceive themselves as susceptible to acquiring HIV. Knowledge can be very life threatening at times that a person with it will feel at risk of becoming a victim because they are aware of the aftermath of their actions if they do not pay attention to their behavior. In this case, a person will be afraid of engaging in any risky sexual behaviors because of the costs attached. However, a person who has shallow or no knowledge about HST may not see themselves at risk because they do not know the costs attached to being unsure of their HIV sero-status and psychological wellbeing.

The above findings are in contrast with a Ghanian study where Ghanian adolescents had a negative impression of their risk of contracting HIV, in other words, they did not find themselves to be at risk. This can be attributed to the kind of behaviors they were engaged in or even ignorance. The findings further suggest that 16% of the male adolescents and 15% of the females thought they were at risk. This represents a small number of adolescents that find themselves susceptible to contracting HIV [22]. A surprising finding also indicates that individuals who felt more susceptible to contracting HIV, 70% males and 59% of the older adolescents, were more engaged in sexual activity [22]. The findings imply that older teenagers compared to those between the ages of 10 and 14, were more likely to believe they were at danger of obtaining HIV. In order to lower their chances of catching HIV, some, almost 57% of females and 47% of males, said they avoided having intercourse, while others, 12% of females and 20% of males, said they used condoms [22].

It was also hypothesized that knowledge of HST is significantly related to individual perceived benefits. The findings confirm that HST knowledge has a positive significant relationship with individual perceived benefits of using HST whereby, possession or acquisition of key information regarding HST use is most likely going to positively increase the use of the intervention. Similarly, an employee, after realizing that HST outweighs any other options and, most importantly, with the added benefits such as privacy, convenience, time saving and easy interpretation among so much more; such an individual is most likely to use the method for he/she had already been equipped with the necessary knowledge and information pertaining HST.

Contrary to the study findings, a study in Kenya conducted by Olakunde and colleagues [23] among young adults found that concerns and misconceptions relating to HST (inaccurate results, harmful, ill-omened) were reasons for non-use of HST. In fact, young adults who stated that they had heard of HST, were, prior to the study, also significantly less likely to be willing to use HST because of their prior knowledge of self-test kits. Participants with the lowest levels of education were also less likely to endorse HST [23]. Students in Uganda who had ever tested for HIV and were sexually active, indicated the need for frequent HIV testing and chose to use self-test kits because they (kits) were practical, simple to use, and guaranteed privacy [24].

It was also hypothesized that HST knowledge is significantly related to individual perceived barriers. The findings indicate that HST knowledge has a significant positive relationship with individual perceived barriers. The results indicate that employees that possess HST knowledge have higher chances of perceiving HST as a barrier. In other words, the knowledge of HST makes them much more worried to attempt and use the service. This could be attributed to the fear that comes should the results turn out negative or, from experiences with people that have in the past succumbed to HIV, hence posing as a barrier to HST. Men, in most cases, have been found unreceptive towards HST due to busy work schedules and demands from aspects of life hence finding HIV testing not only a waste of time, but also, in some cases, taken for granted; they imagine that a negative HIV test result for their spouses who visit health centres during antenatal care [25], implies that they too are negative hence seeing no need of going for HIV testing [26].

It was hypothesized that individual perceived susceptibility positively relates and predict HST use. The study findings indicate that individual’s perceived susceptibility has a significant positive relationship with HST use and predicts HST use by 14%, that is, employees who find themselves highly susceptible to acquire HIV, are more likely going to be drawn towards undertaking HST to ensure they are always safe. In this study, 90% of the employees found themselves to be highly susceptible to contracting HIV depending on either the partners’ behavior or their own. Therefore, this led to 87% of the employees to feel inclined toward using HST if provided at work given the benefits that come along with it.

The findings concur with those of Magno and Castellos who discovered that numerous factors, including STIs, alcohol usage, drug injection, and several-sexual-partners, affect young women’s susceptibility to HIV [27]. The risk perception and willingness to test for HIV were higher among young women who engage in these high-risk behaviors. Finally, students in Uganda who were sexually active chose to use self-test kits because they were practical, simple to use, and guaranteed privacy [24]. However, contrarily, participants in Kenya were considerably less likely to utilize an oral HST kit if they had recently engaged in high-risk sexual behaviors [23].

It was also hypothesized that individual perceived benefits significantly relate and predict HST use among unskilled workers. The findings indicate that individual perceive benefits have a positive significant relationship with HST use and perceived individual benefits stood out with a prediction rate of 44.2% on HST use. Employees who knew about the benefits that result from HST were more prepared to participate in HST since the benefits outweighed the cons. Therefore, to increase HST especially in a busy working environment, benefits pertaining a particular health intervention must be laid out so that respondents get a chance to weigh the risks and benefits of such a practice.

Similarly, self-testing willingness appeared to be mostly motivated by the need to obtain convenient and private information about one’s HIV status [23]. In a study of key HIV stakeholders, HST was viewed as a method of addressing persons who are underserved or unknown. For instance, taking a test in a health centre would not be appropriate in a professional workplace where everyone wears business attire. Such a group is probably going to adhere to the new HST trend [26]. Results from the HST were seen to be more trustworthy than those obtained from regular HIV testing at clinics. Over 80% of Ugandan fishermen who participated in a study of men in Sub-Saharan Africa agreed with the HST findings [28].

It was finally hypothesized that individual perceived barriers significantly relate and predict HST use among unskilled employees. The findings further confirm that individual perceived barriers do have a positive significant relationship with HST use and a prediction level of 4.8% that is, 64% of the employees who perceived other underlying barriers to HST also strongly agreed on using it in other words, be it that HST would pose some unwanted challenges to some individuals such as lack of counselling before and after services, the credibility of results and falsification among others, this did not however limit them from wanting to use the method if provided. Hence affirming the importance of HST.

Equally to the findings, a study in Uganda found out that, men’s non-engagement in HIV testing programs has often been attributed to work commitments and a lack of time [29]. However, Muwanguzi and colleagues confirmed that accessibility to HIV testing services was a motivator for workplace-based HIV self-testing [14]. Self-testing for HIV at work could be a potential solution to the problems with facility-based testing that are now present, such as long wait times, big lineups, and a lack of counsellors [30]. Unfortunately, the difficulty in connecting to care after HIV self-testing still exists [31]. Oduetse and colleagues suggest providing follow-up assistance to everyone who purchases test kits to strengthen the connection to post-test services [32].

## Conclusion

Making individuals aware of the benefits of HST use and equipping them with sufficient knowledge pertaining the desired action would go a long way in curbing down the indulgence in risky sexual behaviors and decrease the spread of HIV especially in populations with limited access to health care.

## Data Availability

Data is available

## Acknowledgements

Research reported in this publication was supported by the Fogarty International Center, National Institute of Alcohol Abuse and Alcoholism, National Institute of Mental Health, of the National Institutes of Health under Award Number D43 TW011304. The content is solely the responsibility of the authors and does not necessarily represent the official views of the National Institutes of Health. In a unique way, we do recognize the Department of Educational, Social and Organizational Psychology for having been a safe harbour and source of wisdom during the entire process and finally the research assistants who made this work come to life.

## Author contributions

**Conceptualization:** Gerald Mukisa Nsereko, Martin Mabunda Baluku, Khamis Musanje, Loyce Kobusingye, Joanita Nangendo

**Formal analysis:** Gerald Mukisa Nsereko, Simon Nantamu, Martin Mabunda Baluku

**Funding acquisition:** Gerald Mukisa Nsereko

**Investigation:** Gerald Mukisa Nsereko

**Methodology:** Gerald Mukisa Nsereko, Martin Mabunda Baluku, Khamis Musanje

**Project administration:** Gerald Mukisa Nsereko, Ayesiga Eron Robinah

**Resources:** Gerald Mukisa Nsereko

**Supervision:** Loyce Kiiza Kobusingye, Martin Mabunda Baluku Khamis Musanje

**Validation:** Gerald Mukisa Nsereko, Loyce Kiiza Kobusingye, Martin Mabunda Baluku Khamis Musanje

**Visualization:** Gerald Mukisa Nsereko, Loyce Kiiza Kobusingye, Martin Mabunda Baluku Khamis Musanje

**Writing – original draft:** Gerald Mukisa Nsereko, Khamis Musanje, Martin Mabunda Baluku

**Writing – review & editing:** Gerald Mukisa Nsereko, Martin Mabunda Baluku, Khamis Musanje, Loyce Kiiza Kobusingye, Simon Nantamu, Joanita Nangendo, and Peter Nsubuga.

